# Gene-Based Rare Variant Burden Analyses Across Biobanks Identify Novel High-Risk Genes for Thoracic Aortic Disease

**DOI:** 10.64898/2026.09.16.26363254

**Authors:** David R. Murdock, Pujun Guan, Dongchuan Guo, Francisca Bermudez, John S. DePaolo, John Cabot, Sumaiya Nazeen, Habib Nasir, Rajat M. Gupta, Alok Jha, John Elefteriades, Bobbi McGivern, Kirsty McWalter, Samantha K. Anderson, Carolyn Jones, Julie A. Lynch, Kyong-Mi Chang, Philip S. Tsao, VA Million Veteran Program, Penn Medicine BioBank, Scott M. Damrauer, Han Chen, Dianna M. Milewicz

**Affiliations:** Division of Medical Genetics, Department of Internal Medicine, McGovern Medical School, The University of Texas Health Science Center at Houston (UTHealth), Houston, TX, USA; Department of Surgery, Perelman School of Medicine, University of Pennsylvania, Philadelphia, PA, USA; Division of Vascular Surgery, Department of Surgery, Stanford University School of Medicine, Stanford, CA, USA; Department of Biomedical Informatics, Harvard Medical School, Boston, MA, USA; Division of Genetics, Brigham and Women’s Hospital and Harvard Medical School, Boston, MA, USA; Broad Institute of MIT and Harvard, Cambridge, MA, USA; Division of Cardiovascular Medicine, Brigham and Women’s Hospital and Harvard Medical School, Boston, MA, USA; Center for Neurogenetics, Weill Cornell Medicine, New York, NY, USA; Aortic Institute at Yale-New Haven Hospital, Yale University School of Medicine, New Haven, CT, USA; GeneDx, LLC, Gaithersburg, MD, USA; Department of Pediatrics, Rush University Medical Center, Chicago, IL, USA; VA Informatics and Computing Infrastructure (VINCI), Salt Lake City VA, Salt Lake City, UT, USA; Department of Internal Medicine, Epidemiology, University of Utah School of Medicine, Salt Lake City, UT, USA; Corporal Michael J. Crescenz VA Medical Center, Philadelphia, PA, USA; Department of Medicine, University of Pennsylvania Perelman School of Medicine, Philadelphia, PA, USA; Department of Medicine, Stanford University School of Medicine, Stanford, CA, USA; VA Palo Alto Health Care System, Palo Alto, CA, USA; Department of Epidemiology, The University of Texas Health Science Center at Houston (UTHealth), Houston, TX, USA

## Abstract

**Background:** Thoracic aortic aneurysms enlarge silently and can cause fatal aortic dissection without timely surgical repair, underscoring the need for improved approaches to identify individuals at high risk. Rare pathogenic variants in established heritable thoracic aortic disease (HTAD) genes explain only a subset of familial and fewer nonfamilial thoracic aortic disease (TAD) cases.

**Methods:** We performed phenotype-stratified, genome-wide, gene-based rare-variant burden analyses of ultrarare damaging missense and predicted loss-of-function variants. Primary analyses focused on aortic dissection, thoracic aortic aneurysm requiring surgical repair, and their combined phenotype. Broader thoracic aortic aneurysm (TAA) was evaluated as a secondary phenotype. Discovery analyses were conducted in the UK Biobank and All of Us, followed by independent replication in the Penn Medicine BioBank, Mass General Brigham Biobank, and Million Veteran Program. Discovery and replication results were subsequently combined in an overall fixed-effect, inverse-variance-weighted meta-analysis across up to five biobanks. Implicated genes were further evaluated in additional clinically ascertained TAD cohorts and using single-cell transcriptomic data from human thoracic aortic tissue.

**Results:** Discovery analyses identified 80 genes reaching study-wide significance across the prespecified TAD phenotypes. These included six established and two putative HTAD genes. Fourteen genes demonstrated independent replication support and reached study-wide significance in the overall meta-analysis across up to five biobanks, which included more than 10,000 cases and 880,000 controls. The eight novel candidate genes among these were *FNDC3B, ROCK1, URM1, SLFN11, ENPP1, CLEC16A, CREM,* and *VCAN*. Associations were strongest for dissection and TAA requiring surgical repair. Four novel associations were driven exclusively by missense variants. *FNDC3B* was observed in a family with HTAD, while additional variants were identified primarily in sporadic dissection or aortic surgery cohorts, suggesting that other genetic or physiologic factors may influence penetrance. The implicated genes showed cell-type-specific expression patterns in human thoracic aortic tissue.

**Conclusions:** These findings expand the genetic architecture of TAD by identifying eight novel candidate genes and demonstrate the utility of phenotype-stratified rare variant burden analyses across large biobanks for gene discovery.

## Introduction

Thoracic aortic disease (TAD) is a major cause of morbidity and mortality^1^. Thoracic aortic aneurysms involving the proximal aorta enlarge asymptomatically over time and may progress to acute type A aortic dissection, a frequently fatal event if not promptly treated^2^. Less common type B dissections initiate in the descending thoracic aorta with little to no prior aortic enlargement^2^. Surgical repair of a proximal aneurysm is typically recommended when the aortic diameter reaches ≥5 cm^2^; however, 60% of patients presenting with acute type A dissections have diameters smaller than 5.5 cm, including cases with no enlargement^3^. These observations underscore the limitations of diameter-based risk stratification and highlight the need for improved approaches to identify individuals at high risk for dissection.

Hypertension and genetic variants are major risk factors for TAD, and genetic risk varies across allele frequencies. Genome-wide association studies (GWAS) have identified common variants associated with TAD at multiple risk loci, including variants in *FBN1* (MIM: 134797), the gene associated with Marfan syndrome (MIM: 154700)^2,4^. At the other end of the spectrum, rare variants that disrupt the coding regions of genes can confer a highly penetrant risk for TAD, with up to 20% of patients having a similarly affected family member, termed heritable TAD (HTAD)^5^. While Mendelian approaches have identified 11 confirmed and additional putative HTAD genes, including *FBN1*, pathogenic variants (PVs) in these genes account for only 30% of HTAD families and 10% of early-onset dissections, indicating that additional high-risk genetic variants remain to be discovered^5,6^. Recent work shows that genetic testing achieves high diagnostic yield in selected patients using clinical criteria, including early age of onset, syndromic features, family history, and absence of hypertension, but may not explain the broader population in whom genetic variants still contribute to disease risk^7^. Similar gaps have been observed in other cardiovascular diseases, such as heart failure, where common variants identified by GWAS and rare variants in established cardiomyopathy genes account for only a fraction of heritability, suggesting that additional high-risk genetic contributors remain to be identified^8^.

Single-variant tests commonly used in GWAS are underpowered to detect associations of rare alleles, but aggregating rare variants across a gene can overcome this limitation^9,10^. While such methods have successfully identified genes enriched for loss-of-function (LOF) variants, detecting disease associations driven by rare missense variation has proven more challenging^11,12^. Improving the genome-wide detection of high-impact rare variants across the mutational spectrum is therefore critical for advancing genetic discovery in TAD and other cardiovascular diseases.

In this study, we developed and applied a phenotype-stratified framework for gene-based rare variant burden testing. This approach complements common-variant GWAS and family-based gene discovery by testing whether distinct ultrarare damaging variants within the same gene are enriched across unrelated individuals with a shared phenotype. Primary analyses focused on severe TAD phenotypes, including dissection and thoracic aortic aneurysm (TAA) requiring surgical repair, based on prior work suggesting greater enrichment for high-risk rare variants than in broader TAA^13^. The framework incorporated gnomAD frequency, REVEL, and AlphaMissense thresholds that we previously derived and validated across independent cohorts to identify rare, damaging variants that increase aortic dissection risk^13–16^. Using harmonized data from five biobanks comprising more than 10,000 TAD cases and 880,000 controls, we performed discovery analyses in the UK Biobank and All of Us, followed by independent replication in the Penn Medicine Biobank, Mass General Brigham Biobank, and Million Veteran Program. Results were then combined in an overall meta-analysis across up to five biobanks. Implicated genes were further assessed using single-cell transcriptomic data from normal and diseased human thoracic aortas, as well as additional clinically ascertained cohorts. This approach identified established HTAD genes and eight novel candidate genes, including associations driven solely by damaging missense variants.

## Methods

### Biobank Cohorts

This study was approved by the institutional review board (IRB) at the University of Texas Health Science Center in Houston (UTHealth) in accordance with the ethical standards of the Declaration of Helsinki. Discovery analyses were performed in the UK Biobank (UKB) and the All of Us (AoU) Research Program^17,18^. Across these cohorts, 4,889 individuals had a diagnosis of dissection or TAA, and 757,960 individuals were classified as controls. The prespecified primary phenotypes included 1,063 dissection cases, 418 cases of thoracic aortic aneurysm undergoing surgical repair (TAA-surgery), and 1,481 individuals with either dissection or TAA-surgery. Replication analyses were conducted in three independent cohorts: the Penn Medicine Biobank (PMBB), Mass General Brigham Biobank (MGBB), and the VA Million Veteran Program (MVP), contributing an additional 5,412 TAD cases and 123,004 controls^4,19,20^. Cohort characteristics are detailed in the Supplemental Methods and Table S1.

### TAD Phenotype Definitions

TAD phenotypes were defined using International Classification of Diseases (ICD) and procedure codes (Table S2), adapted from Klarin *et al.*^4^. For the discovery analyses in UKB and AoU, we prespecified primary and secondary TAD phenotypes. Primary phenotypes comprised clinically severe presentations, including aortic dissection (Dissection), TAA-surgery, and a combined phenotype including all individuals with either dissection or TAA-surgery (Dissection + TAA-surgery). Dissection and TAA-surgery cases were mutually exclusive. Secondary analyses were performed in individuals with thoracic aortic aneurysm (TAA) to assess the burden of rare variants across the broader spectrum of aneurysm presentations.

Controls were defined as individuals without any TAD case codes and without diagnostic codes for aortic disease, congenital malformations, deformations, or chromosomal abnormalities, or established HTAD (e.g., Marfan syndrome). To ensure consistent phenotype definitions across cohorts, all ICD codes were harmonized to the UKB format, which omits decimal subcodes.

### Additional Clinically Ascertained Cohorts

Independent clinically ascertained cohorts were evaluated to assess candidate genes: the UTHealth HTAD cohort (437 probands from families with ≥2 members with TAD), the Early-Onset Sporadic Thoracic Aortic Dissection (ESTAD) cohort (551 individuals aged ≤60 years), and the Yale aortic surgery cohort (1,750 adults undergoing thoracic aortic aneurysm or dissection repair). Exome or genome sequencing was performed in each cohort. A clinically ascertained trio identified through physician-ordered sequencing was also included. Additional details are provided in the Supplemental Methods.

### Variant Annotation and Filtering

Sequencing data were processed using standardized pipelines with rigorous quality control measures, as detailed in the Supplemental Methods. For missense variant burden analyses, we applied previously derived thresholds for gnomAD minor allele frequency (MAF), REVEL, and AlphaMissense^14–16^. These thresholds were derived in the PMBB using a data-driven cutpoint approach based on the Youden index, internally validated by bootstrapping, and subsequently validated in independent cohorts^13^. Missense variants were therefore restricted to MAF < 8.16 × 10⁻⁶, REVEL > 0.649, and AlphaMissense > 0.2543 across 19,252 protein-coding genes^13^. Predicted loss-of-function (LOF) variant analyses were similarly restricted to rare variants (MAF < 8.16 × 10⁻⁶) with stop-gain, canonical splice-site, or frameshift consequences in 3,951 genes defined as having a Probability of Loss-of-function Intolerance (pLI) score ≥ 0.9 or high arterial expression and ≤ 30 LOF alleles in gnomAD. The combined burden of missense and predicted LOF variants was also evaluated in these 3,951 genes^14^.

### Rare Variant Burden Analysis

Gene-based rare-variant burden testing was performed separately in UKB and AoU using Firth’s penalized logistic regression, modeling case-control status as a function of rare-variant carrier status with covariates for age, sex, and the first four (UKB) or five (AoU) genetic principal components^21^. Related individuals (kinship coefficient Φ > 0.1) and individuals with missing covariates or sex discordance were excluded, with cases preferentially retained. For autosomal genes, heterozygous and homozygous carriers were treated equivalently, and for X-linked genes, hemizygous males and heterozygous or homozygous females were considered carriers. For each gene, logistic regression was performed separately for each combination of variant class (missense, LOF, or combined missense + LOF) and TAD phenotype group (e.g., aortic dissection). Only gene–variant class–phenotype combinations with at least one case carrier in both UKB and AoU were included in the discovery meta-analysis.

Meta-analysis results from UKB and AoU were combined using an inverse-variance-weighted fixed-effects model to obtain an overall estimate under the assumption of a common genetic effect across cohorts, with heterogeneity assessed using Cochran’s Q test^22,23^. Study-wide significance thresholds were defined using a Bonferroni correction for the number of genes tested (α = 0.05 / 19,252 = 2.6 × 10⁻⁶ for missense; α = 0.05 / 3,951 = 1.3 × 10⁻⁵ for LOF and combined analyses) using two-sided P values. For each gene, only the most significant variant class–TAD group association reaching study-wide significance was carried forward for replication testing. Because of data-use and disclosure-control policies across the contributing biobanks, cohort-specific numbers of qualifying variants, carrier counts, case-control carrier breakdowns, and allele-level summaries that could disclose or permit derivation of restricted participant counts were not publicly reported. Aggregate carrier and non-carrier counts are provided in the supplemental tables where permitted.

### Independent Replication and Five-Cohort Meta-Analysis

Genes that reached study-wide significance in the UKB + AoU discovery meta-analysis were carried forward for replication in three independent biobank cohorts (PMBB, MGBB, and MVP). Analyses were performed using the same statistical framework as in the discovery phase. For replication cohorts, analyses were conducted using a harmonized thoracic aortic aneurysm and/or dissection (“TAD”) case definition to ensure consistent phenotype application, using the same diagnostic and procedure codes as in discovery (Table S2). Replication cohorts without any case carriers for a given gene were excluded. Independent replication support was defined as (i) nominal significance (P < 0.05) in at least one replication cohort and (ii) directional consistency of the effect estimate with the discovery association.

Logistic regression results from up to five biobanks were combined using inverse-variance-weighted fixed-effects meta-analysis to obtain overall summary estimates, as in the discovery phase. Because cohort-specific differences in phenotype definitions, ancestry, and other factors could introduce heterogeneity, a DerSimonian–Laird random-effects meta-analysis was also performed as a sensitivity analysis^24^. Genes were considered to meet the prespecified cross-cohort validation criteria if they demonstrated independent replication support and reached Bonferroni significance in the combined five-cohort meta-analysis.

As an additional sensitivity analysis, hypertension was defined using ICD code I10 and included as a covariate in the UKB and AoU discovery analyses for genes meeting the cross-cohort validation criteria. Diagnosis-based hypertension was used because measured systolic and diastolic blood pressure and antihypertensive medication exposure were not uniformly available or harmonized across cohorts. To assess potential effect modification by sex, sex-stratified burden analyses and gene-by-sex interaction testing were also performed in UKB and AoU for these genes.

## Results

### UK Biobank and All of Us Discovery Analysis

In the UKB and AoU discovery meta-analysis, comprising 4,889 TAD cases and 757,960 controls, genome-wide rare variant burden testing identified 80 genes with a study-wide significant association with one or more TAD phenotypes (Table S3; Figure 2). These associations had large estimated effect sizes, with a geometric mean odds ratio (OR) of approximately 25. Six of the 11 established HTAD genes were identified: *FBN1*, *ACTA2* (MIM: 102620), *SMAD3* (MIM: 603109), *TGFBR2* (MIM: 190182), *COL3A1* (MIM: 120180), and *MYLK* (MIM: 600922), as well as putative genes *FLNA* (MIM: 300017) and *PKD1* (MIM: 601313), supporting the validity of this approach^6^. Burden of only missense variants in *FBN1* was significantly increased in dissection + TAA-surgery cases (P = 4.1 × 10^-22^; OR 8.9, 95% CI 5.7–13.8), with LOF variants showing an even greater burden (P = 3.5 × 10^-71^; OR = 413.0, 95% CI 213.1–800.6)^25,26^. Missense variants in *ACTA2* cause non-syndromic HTAD (MIM: 611788), and the burden of both missense variants only (P = 3.0 × 10^-23^; OR = 28.3, 95% CI 14.6–54.7) and missense + LOF rare variants (P = 4.1 × 10^-25^; OR = 27.6, 95% CI 14.7–51.6) were also significantly increased among dissection + TAA-surgery cases^27,28^. Rare variants in *TGFBR2*, *SMAD3, COL3A1*, and *MYLK* likewise achieved study-wide significance, consistent with their established TAD risk in Loeys–Dietz syndrome 2 and 3 (MIM: 603109, 610168), vascular Ehlers–Danlos syndrome (MIM: 130050), and non-syndromic HTAD (MIM: 613780), respectively^5,29,30^. *PKD1* and *FLNA* also reached study-wide significance, strengthening evidence for their proposed role in TAD^5,6^. Notably, all established and putative HTAD genes showed a significantly increased burden of rare variants in genome-wide analyses, despite these associations being driven by relatively few affected carriers per gene. Other HTAD genes showed an increased burden of rare variants in TAD cases but did not reach significance, including *LOX* (MIM: 153455) and *LTBP3* (MIM: 602090)^31,32^.

Seventy-two additional candidate genes reached study-wide significance in the discovery burden meta-analyses, with the majority having the greatest burden of rare variants in either the dissection or dissection + TAA-surgery phenotypes. The burden analyses were significant for only missense variants in 46 genes, LOF in 16 genes, and missense + LOF in 10 genes (Table S3).

### Independent Replication and Five-Biobank Meta-Analysis

Genes reaching study-wide significance in discovery were further assessed in the PMBB (1,657 cases and 40,231 controls), MGBB (426 cases and 8,980 controls), and MVP (3,329 cases and 73,793 controls) using the same variant filters and analytical framework (Figure 1). For each gene, the burden model corresponding to the variant class with the strongest discovery-phase association was carried forward. Replication analyses evaluated the corresponding variant class using a harmonized case definition that captured thoracic aortic aneurysm and/or dissection (“TAD”) to ensure consistent case definitions across replication biobanks, and used the same diagnostic and procedure codes as in discovery (Table S2). Results from these cohorts were combined with those from the UKB and AoU in a meta-analysis of up to five biobanks using an inverse-variance-weighted fixed-effects model^23^.

**Figure 1.**
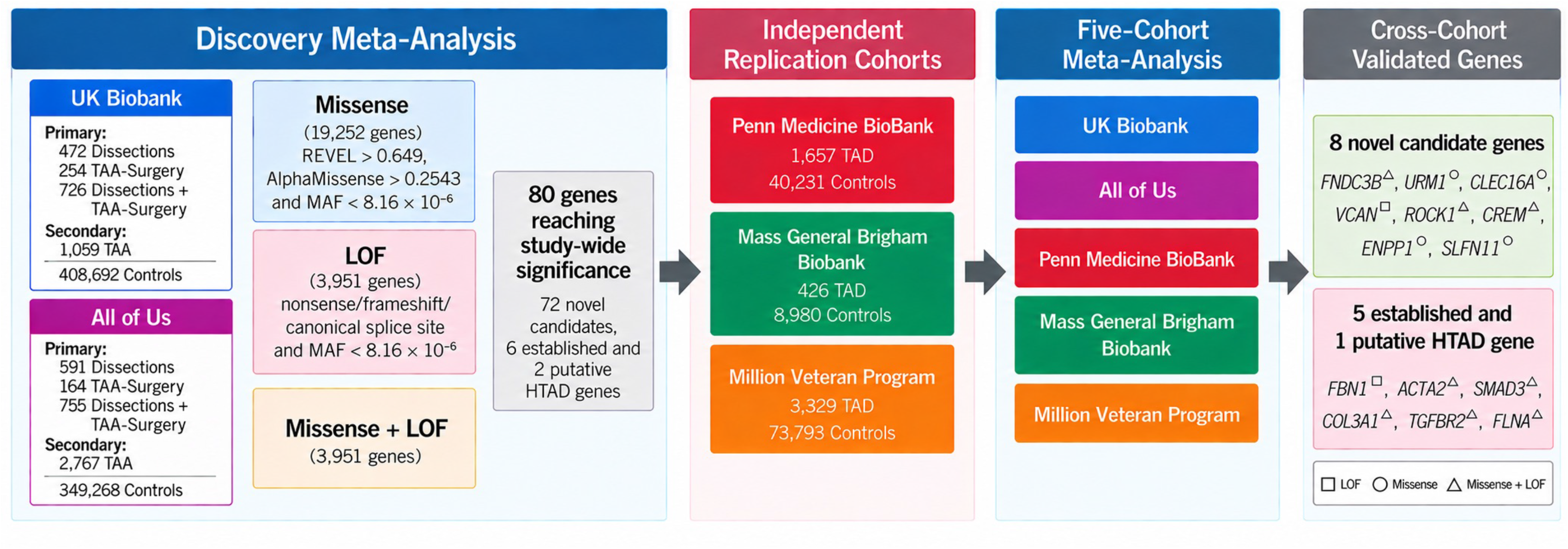
Study design and meta-analytic framework for rare variant burden testing in TAD. Rare variant burden testing was performed in the UK Biobank and All of Us cohorts across missense, loss-of-function (LOF), and combined missense + LOF models, followed by fixed-effect meta-analysis. Eighty genes reached study-wide significance (6 established and 2 putative HTAD genes and 72 novel candidates) and were carried forward for independent replication in the Penn Medicine BioBank (PMBB), Mass General Brigham Biobank (MGBB), and Million Veteran Program (MVP). Independent replication support was defined as nominal significance (P < 0.05) in at least one replication dataset with a concordant effect direction. Results from the discovery and replication cohorts were subsequently combined in a five-cohort meta-analysis. Fourteen genes met the cross-cohort validation criteria in the combined meta-analysis of up to five cohorts, including eight novel candidate genes, five established HTAD genes, and one putative HTAD gene. Superscript symbols indicate the variant class for each gene’s most significant discovery-phase phenotype–variant class association: loss-of-function (squares), missense (circles), or combined missense + loss-of-function (triangles).

**Figure 2.**
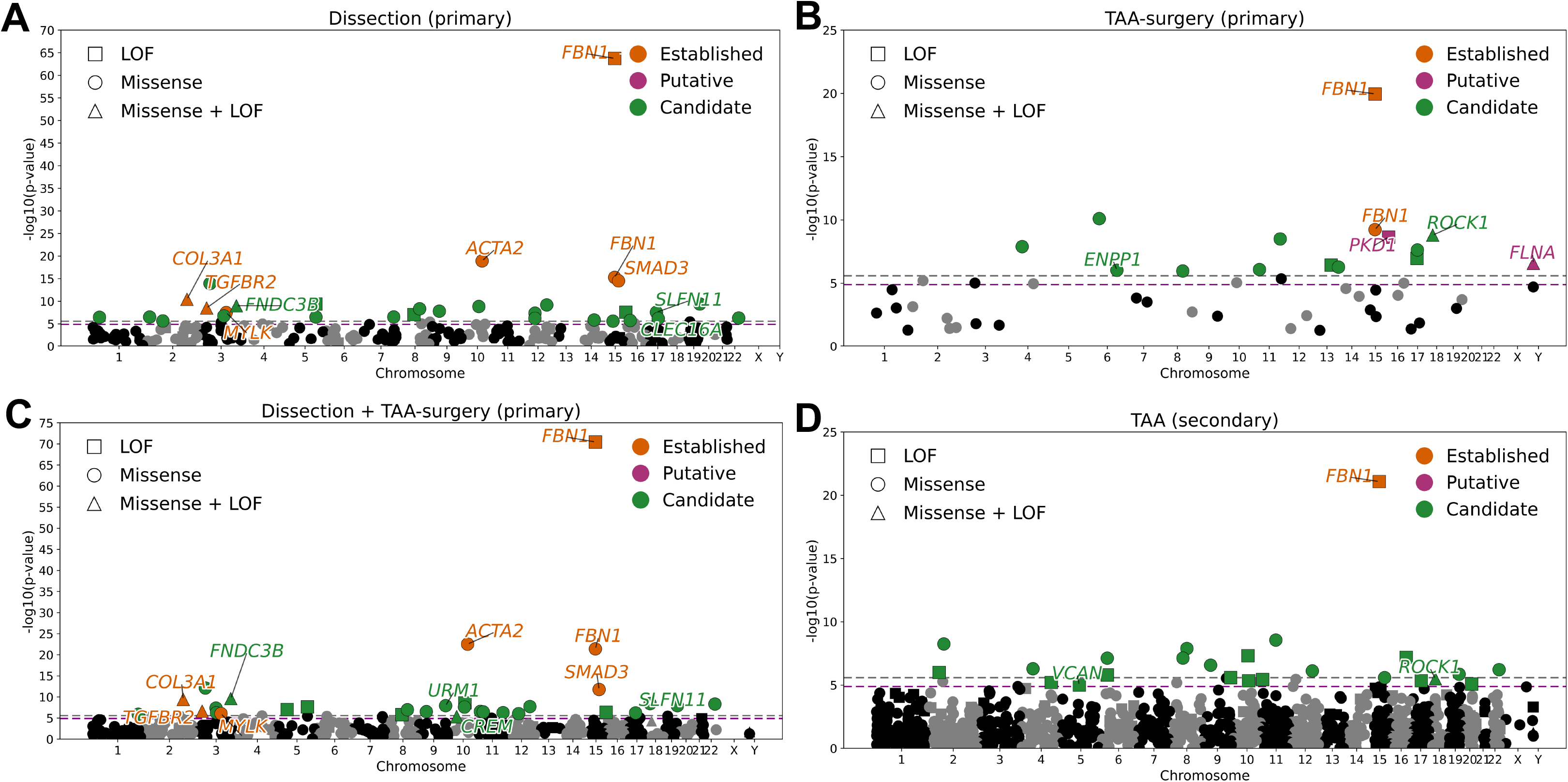
Discovery meta-analysis of rare variant burden across TAD phenotype groups. **A–D** UK Biobank + All of Us discovery meta-analysis results for Dissection, TAA-surgery, Dissection + TAA-surgery, and TAA. Panels A–C correspond to primary analyses of dissection and TAA-surgery phenotypes, including their combined analysis, whereas panel D shows the secondary analysis of TAA cases alone. For genes reaching study-wide significance, significant missense and loss-of-function (LOF) associations are shown separately; the combined missense + LOF association is shown only when it is the sole variant class reaching study-wide significance for that gene. Shapes indicate LOF (squares), missense (circles), or combined missense + LOF (triangles). Colors denote established HTAD genes (orange), putative HTAD genes (purple), and candidate genes (green). Dashed lines indicate Bonferroni-corrected significance thresholds (grey, P = 2.6 × 10⁻⁶ for missense; purple, P = 1.3 × 10⁻⁵ for LOF and combined missense + LOF analyses). Candidate gene labels are shown only for associations that met the cross-cohort validation criteria. Full results are provided in Table S3.

Of the 80 genes significant in the discovery-phase meta-analysis, 14 demonstrated independent replication support and reached study-wide significance after Bonferroni correction in the combined five-biobank meta-analysis (Table 1, Table S4). Of these, eight were novel candidate genes: *FNDC3B*, *ROCK1*, *URM1*, *SLFN11*, *ENPP1*, *CLEC16A*, *CREM*, and *VCAN* (Figure 3). The remaining six were established or putative HTAD genes, *FBN1*, *ACTA2*, *SMAD3*, *COL3A1*, *TGFBR2*, and *FLNA*, three of which showed study-wide significant associations in missense-only analyses (Figure S1). Moderate between-cohort heterogeneity was observed for some genes, most likely due to variation in sample sizes, phenotype distributions, and the stochastic presence of ultra-rare variants across biobanks. However, the DerSimonian and Laird random-effects estimates were broadly consistent with the fixed-effect results, supporting the robustness of these signals and indicating that the associations persisted after accounting for cohort-specific differences (Table S5)^23,24^. Sensitivity analyses additionally adjusting for hypertension produced broadly similar effect estimates for the 14 genes meeting cross-cohort validation criteria (Table S6). Sex-specific burden estimates were available in UKB and AoU for 7 genes in females and 8 genes in males. The available estimates were generally directionally consistent with the primary associations, but sparse carrier counts precluded definitive conclusions regarding sex-specific effects (Table S7). *FBN1* showed nominal evidence of a stronger burden association in females than males in the meta-analysis (interaction OR, 0.21 [95% CI, 0.05–0.87]; *P* = 0.032), although the interaction was significant only in UKB. No significant interactions were observed for *SMAD3*, *FLNA*, or *VCAN*, whereas low carrier counts limited analyses of the remaining genes.

**Figure 3.**
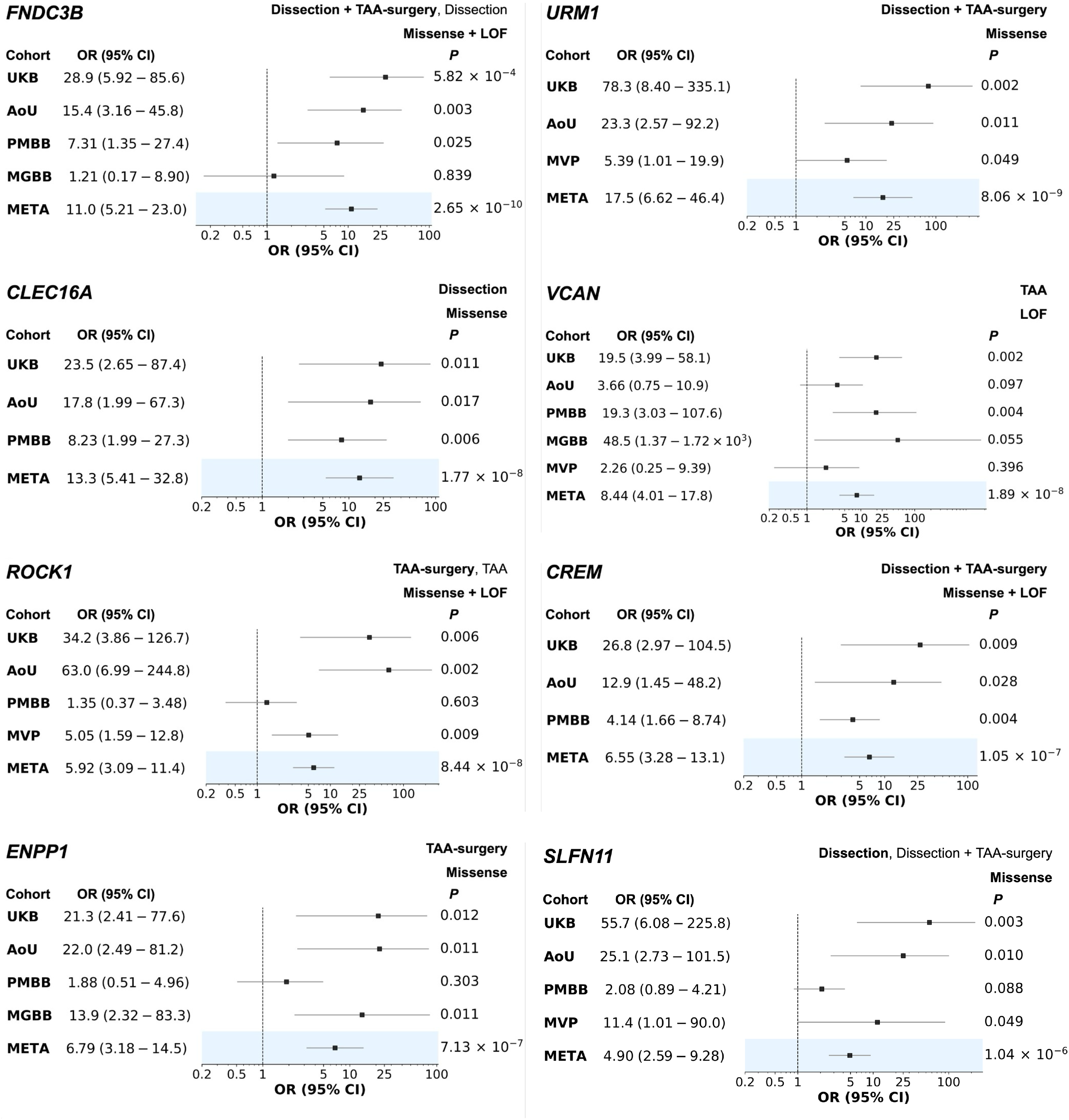
Cross-cohort validation of novel candidate genes associated with TAD. Cohort-specific and fixed-effect inverse-variance weighted meta-analytic odds ratios (ORs) with 95% confidence intervals and *P* values are shown for the eight novel candidate genes that met the cross-cohort validation criteria. Meta-analyses included up to five cohorts: UK Biobank (UKB), All of Us (AoU), Penn Medicine BioBank (PMBB), Mass General Brigham Biobank (MGBB), and Million Veteran Program (MVP). Cohorts without case carriers for a given gene–variant class association were excluded from the corresponding meta-analysis. For each gene, the TAD phenotype(s) and variant class (missense, LOF, or missense + LOF) that reached study-wide significance in the UKB + AoU discovery meta-analysis are indicated. The phenotype–variant class combination carried forward for cross-cohort validation is indicated in **bold**. Primary analyses focused on dissection and TAA-surgery phenotypes, including their combined analysis. *VCAN* was the only gene for which the association was restricted to the secondary TAA phenotype. LOF, loss-of-function. Full results are provided in Table S9.

**Table 1.**
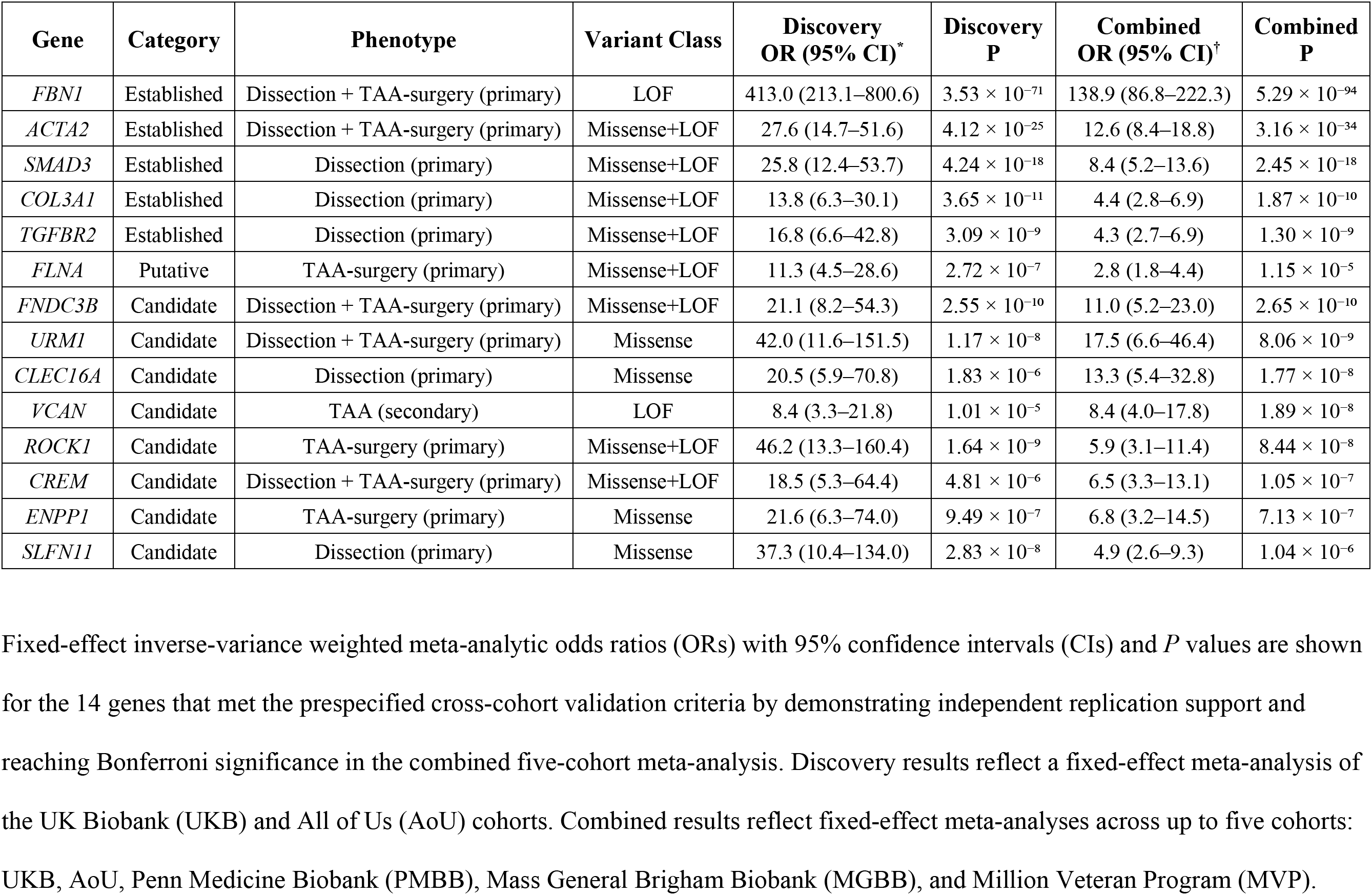
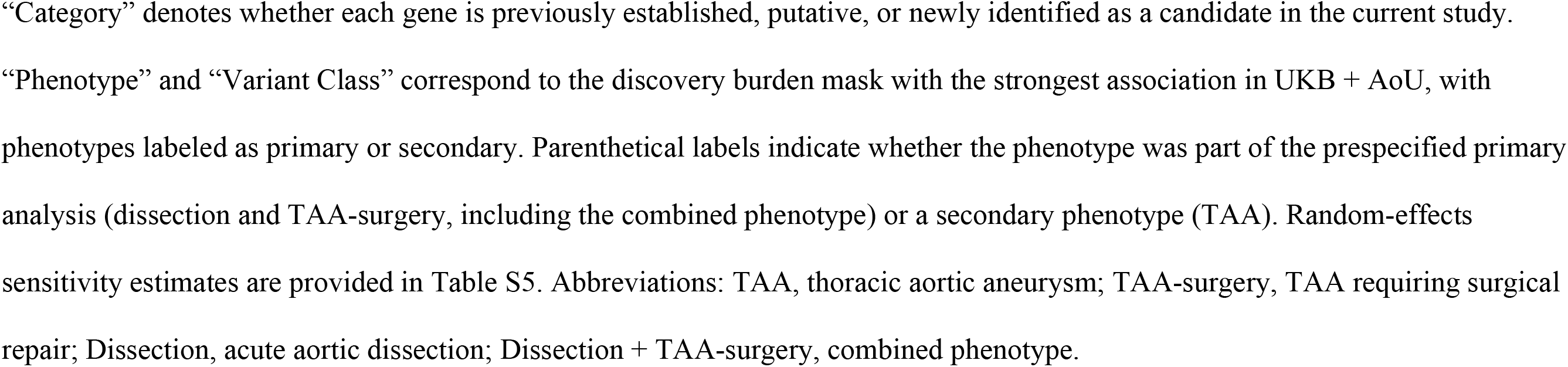
Discovery and cross-cohort validation of rare variant associations with TAD.

For the novel genes, both the variant type and the associated TAD phenotype varied. The most significant burden was identified for *FNDC3B*, *URM1*, and *CLEC16A*, in dissection cases or combined dissection + TAA-surgery cases. In the discovery-phase analysis, these genes showed effect sizes comparable to those of established HTAD genes, with ORs of approximately 20–40 (Table 1, Tables S8–S9). The reduced ORs in the extended meta-analysis are consistent with inflated effect sizes in discovery analyses (i.e., the Winner’s Curse) and are further influenced by the inclusion of smaller, more heterogeneous replication cohorts^33–35^. Four genes (*URM1*, *CLEC16A*, *ENPP1*, and *SLFN11*) showed an increased burden restricted to missense variants, primarily in dissection or dissection + TAA-surgery cases, with *ENPP1* showing a missense-specific association limited to TAA-surgery. *ROCK1* and *CREM* showed increased burden of combined missense and LOF variants, with *ROCK1* associated with TAA and TAA-surgery and *CREM* with dissection + TAA-surgery. *VCAN* showed a LOF-specific burden restricted to TAA cases. Thus, the methods used for these analyses identified novel genes with an increased burden of missense variants, LOF variants, or both.

### Assessment of Novel Genes in Additional TAD Cohorts

We evaluated the eight novel genes for rare variants in unsolved cases in independent TAD cohorts (Table S10). In the UTHealth ESTAD cohort, four LOF variants in *FNDC3B* were identified, and a Fisher’s exact test of burden in age/sex-matched and sequencing-batch-matched controls demonstrated significant enrichment (4/442 cases vs. 0/760 controls; P = 0.018). Three additional *FNDC3B* LOF variants were observed in the Yale TAD cohort of surgically repaired TAA cases and dissections, including a recurrent nonsense variant, c.1453C>T (p.Arg485*), identified in an unrelated ESTAD case. A heterozygous *FNDC3B* frameshift variant, c.3004_3005del (p.Lys1002Glufs*69), was also identified by GeneDx in a family with HTAD; the proband was a child with mild aortic root dilation (z = 2.4), and his father had negative genetic testing and an aortic root replacement in his twenties. A LOF *VCAN* variant, c.221dup (p.Asn74Lysfs*51), was also identified in a Yale TAD case, and an ESTAD case had a *SLFN11* variant, c.2310_2311del (p.Gln771Glyfs*9), that disrupted the C-terminus of the protein.

### Aortic Cell-type Expression of HTAD and Novel TAD-associated Genes

We sought to characterize the expression of genes with an increased burden of rare variants across cell types in the aorta using publicly available human single-cell RNA sequencing data from ascending thoracic aortas (Supplemental Methods)^36,37^. The dataset included transcriptomic data from 5 normal aortas, 8 aortas from patients with TAA undergoing surgical repair, and 6 aortas from patients presenting with type A dissections. Definitive and putative HTAD genes showed distinct cell-type patterns (Figure 4a-c). *ACTA2* was highly expressed in smooth muscle and microvascular populations, and *FLNA* in SMCs, whereas *FBN1* and *COL3A1* were more prominent in fibroblast and mesenchymal progenitor populations, with intermediate levels in SMCs. *SMAD3* showed low-to-moderate abundance across stromal, endothelial, and immune cell populations, while *TGFBR2* was more prominent in endothelial, fibroblast, and mesenchymal progenitor populations, with intermediate levels in SMCs. Among the novel genes, *FNDC3B* was highest in monocytes, with additional signal in macrophages, fibroblasts, and mesenchymal progenitors. *ROCK1* was broadly expressed across mural and immune cell populations, with greater abundance in microvascular mural cells, while *CREM* was highest in T-cells and also detected in NK and myeloid populations. *VCAN* was more abundant in monocytes, with similar levels in SMCs, fibroblasts, and mesenchymal progenitors, whereas *ENPP1* showed low overall abundance with greater expression in SMCs. *URM1*, *CLEC16A*, and *SLFN11* were expressed at generally low levels, with distinct patterns across endothelial, immune, stromal, or Schwann/glial populations. These cell-type patterns were generally similar across control, TAA, and dissection samples, with *VCAN* showing higher expression in TAD aortas.

**Figure 4.**
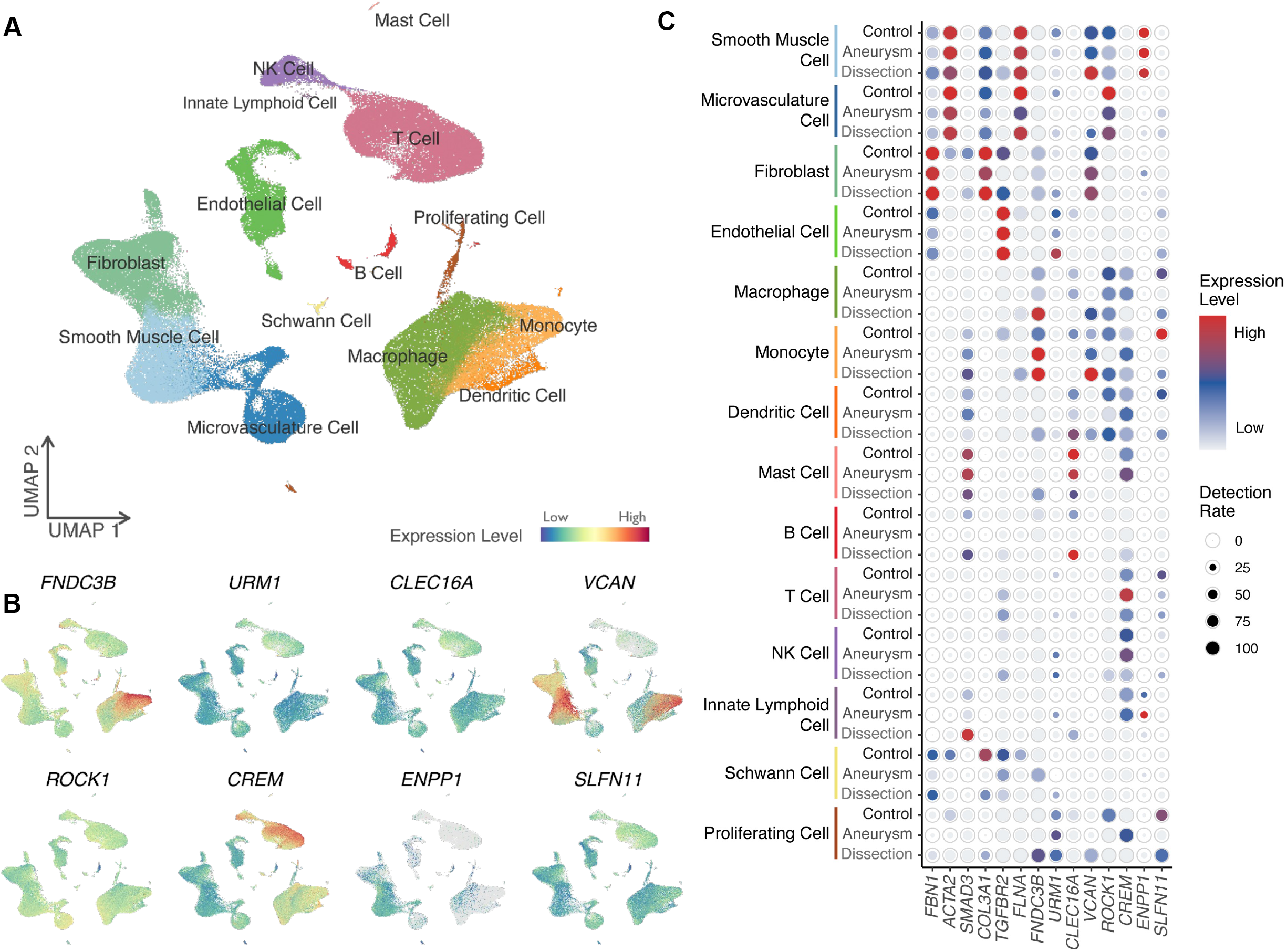
Single-cell expression profiling of novel candidate genes in human aortic tissue. A Uniform Manifold Approximation and Projection (UMAP) visualization showing the major cell populations identified in human aortic tissue, including smooth muscle cells, fibroblasts, endothelial cells, and immune cell subsets. B Feature plots illustrating the expression distribution of the eight novel candidate genes across the UMAP embedding. The color gradient indicates relative expression level (blue: low; red: high). C Dot plot summarizing expression of the novel candidate genes and selected established HTAD genes across cell types and disease states (control, aneurysm, and dissection). Dot size represents the percentage of cells expressing each gene, and color represents the average scaled expression level (blue: low; red: high).

## Discussion

Discovery of rare variants in novel genes by gene-based burden analyses has lagged behind the success of GWAS for common variants, in part because highly penetrant pathogenic alleles are ultrarare and available cohorts are often underpowered to detect them. As large biobanks increasingly provide genomic-level data, the aggregation of rare variants across genes has enabled the detection of high-effect associations, although prior successes have largely focused on LOF variants, with missense-driven associations proving more difficult to identify^11,12^. We therefore applied previously validated thresholds for ultrarare damaging variants to phenotype-stratified gene burden testing across multiple biobanks. This framework recovered established HTAD genes and identified novel high-effect candidates, expanding the genetic architecture of TAD and implicating relevant disease pathways.

The recovery of recognized HTAD genes, including *FBN1*, *ACTA2*, *COL3A1*, *SMAD3*, *TGFBR2*, and *FLNA*, supports this framework’s ability to identify genes harboring high-impact rare variants associated with TAD^5,6^. Notably, *FBN1*, *ACTA2*, and *SMAD3* remained study-wide significant when analyses were restricted to missense variants, demonstrating that stringent prioritization of ultrarare missense variants can identify disease genes without incorporating LOF alleles. *MYLK* and *PKD1* were also identified in the UKB–AoU discovery analysis but did not meet cross-cohort validation criteria. For *MYLK*, pathogenic variants are restricted to the short isoform expressed in aortic tissues, which may reduce the number of informative variants and limit replication across cohorts^38^. The absence of several other established HTAD genes from the discovery results is consistent with gene-specific molecular constraints: *PRKG1* disease risk is driven by a single, recurrent gain-of-function variant^39^; *MYH11* pathogenic variants cluster in a limited region of the coiled-coil domain^40^; and *TGFBR1* pathogenic variants are less frequently observed in population biobanks than *TGFBR2*^13^.

The objective of this study was to identify novel genes harboring variants that increased the risk for TAD, and eight such genes were identified. *URM1*, *CLEC16A*, *ENPP1*, and *SLFN11* showed significance exclusively for missense variant burden. *FNDC3B*, *ROCK1*, and *CREM* were identified through combined missense + LOF burden analyses. *VCAN* was identified only in the LOF analysis. Only rare variants in FNDC3B were observed in HTAD families, suggesting that the newly implicated genes may influence TAD risk through mechanisms distinct from those of highly penetrant HTAD genes, despite comparable effect sizes. Possible explanations include limited power to detect familial segregation or false-positive associations, stochastic factors, and gene-specific mechanisms in which disease risk depends on the genetic or physiological context. Another possibility is that these genes may also increase susceptibility to TAD in the presence of a second “hit”, such as hypertension, the major clinical risk factor for TAD, which is present in >80% of dissection cases^41^. This hypothesis could not be formally evaluated here because the rarity of these variants limited the number of carriers available for stratified analyses by hypertension status. Sensitivity analyses adjusting for hypertension yielded broadly similar effect estimates, but they do not determine whether hypertension modifies penetrance or contributes to disease expression. Notably, individuals with HTAD pathogenic variants develop disease in the absence of hypertension, indicating that elevated blood pressure is not required for TAD onset. Additionally, the rare-variant burden in known and novel genes is higher in dissection cases than in TAA cases, based on our results, suggesting that rare variants in these genes may contribute more to dissection risk than to aneurysm formation alone^13^. Finally, sex-stratified analyses were exploratory because few carriers were available, and the nominal *FBN1* interaction was significant only in UKB.

An important analytic feature of this study is the variant selection strategy. Rather than applying broad rare-variant masks, we restricted analyses to ultrarare alleles enriched for deleteriousness, using thresholds optimized in known HTAD genes^13^. Because these variants are expected to act consistently in a risk-increasing direction, Firth penalized logistic regression provides robust type I error control and interpretable effect-size estimates, whereas kernel-based (e.g., SKAT or Meta-SAIGE) approaches are better suited for mixed-direction effects^21,42–44^. Although this approach is less sensitive to modest or heterogeneous effects, it improves the detection of highly penetrant missense and LOF variants in biobank-scale data. This strategy may also be applicable to other genetically heterogeneous cardiovascular disorders in which rare, deleterious coding variants contribute to disease risk but validated genes explain only a subset of cases, such as dilated cardiomyopathy and Brugada syndrome^45,46^.

Single-cell transcriptomic analyses showed that the novel candidate genes, like definitive and putative HTAD genes, were distributed across multiple aortic cell populations. Several definitive HTAD genes likewise showed substantial expression outside SMCs, underscoring the diverse cellular distribution of genes implicated in TAD. Among the novel candidates, *FNDC3B* and *ROCK1* showed relatively broad cellular distributions, whereas others were more prominent in specific populations, including *CREM* in lymphoid cells and *ENPP1* in SMCs. *VCAN* was the only candidate showing higher expression in TAD aortas. Together, these findings provide cellular context for the genetic associations and suggest that the newly implicated genes may influence TAD through multiple cellular compartments within the aortic wall.

#### Box | Biological Context for Novel Candidate Genes Implicated in TAD

***FNDC3B*** encodes a 132-kDa protein that contains an amino-terminal proline-rich motif, nine fibronectin type III domains, and a transmembrane domain, and is expressed in many tissues. FNDC3B suppresses TGFβ signaling to promote epithelial-to-mesenchymal transition by cancer cells and adipocyte differentiation^47–50^. *Fndc3b^-/-^* mice die shortly after birth of pulmonary disease and have craniosynostosis-like premature ossification of the calvarial bone, similar to cranial abnormalities associated with severe *TGFBR1* and *TGFBR2* pathogenic variants^48,51,52^; 3q26.31 microdeletions involving *FNDC3B* have also been associated with craniofacial anomalies^53^. FNDC3B negatively regulates both differentiation and BMP/Smad signaling pathways that are required for proper calvarial bone development^54^. These observations suggest that loss of *FNDC3B* may alter TGF-β signaling networks relevant to TAD. Further support for *FNDC3B* as a disease-causing gene is based on the identification of *FNDC3B* LOF variants that segregate with disease in HTAD pedigrees and are *de novo* in affected probands from trios (Caluseriu, personal communication).

***VCAN*** (MIM: 118661) encodes versican, a large chondroitin sulfate proteoglycan of the extracellular matrix^55^. *VCAN* has four major isoforms generated by alternative splicing of exons 7 and 8: V0 contains both exons, V1 lacks exon 7, V2 lacks exon 8, and V3 lacks both exons. Three *VCAN* variants identified here in TAD cases occur in exon 7 and are predicted to eliminate V0- and V2-containing transcripts via nonsense-mediated decay (NMD), thereby shifting the isoform balance toward V1 and V3. A variant in an additional case is predicted to disrupt exon 2 splicing and potentially affect all major *VCAN* isoforms via NMD. *VCAN* has been identified as a marker of a fibroblast-like, proliferative SMC population in human TAA, and such SMC phenotypic remodeling may contribute to adaptive responses that maintain aortic wall integrity^56,57^. Importantly, *VCAN* pathogenic variants cause Wagner vitreoretinopathy (MIM: 143200) through isoform imbalance, specifically increased levels of V2 and V3, underscoring the importance of *VCAN* isoform balance in tissue maintenance^55^.

***ROCK1*** encodes Rho-associated coiled-coil containing protein kinase and is highly and selectively expressed in vascular SMCs^49^. ROCK1 is activated by the small GTPase RhoA, forming the RhoA/ROCK signaling pathway, which regulates actin-myosin contractility, stress fiber formation, and focal adhesion assembly. RhoA/ROCK activity is increased early during TAA formation in a mouse model of TAD, and blocking RhoA/ROCK activity increases the incidence of type A aortic dissections, suggesting that this signaling is protective^58^.

***CLEC16A*** contains common variants that have been associated with several autoimmune disorders on GWAS, including juvenile-onset diabetes, lupus, alopecia, and multiple sclerosis^59–62^. The gene encodes an endosomal protein that is critical for mitochondrial function through its regulation of mitophagy^63,64^. Excessive mitochondrial energy workload in aortic SMCs has emerged as a contributor to TAD, and mitophagy is essential for mitochondrial quality control^65,66^.

***ENPP1***, an ectonucleotide pyrophosphatase/phosphodiesterase 1, hydrolyzes extracellular ATP to generate pyrophosphate, an inhibitor of calcification^67^. Biallelic *ENPP1* LOF variants cause generalized arterial calcification of infancy, characterized by calcification by SMCs in the medial layer of arteries^67,68^. ENPP1 is also responsible for the hydrolysis of cGAMP, which suppresses the cGAS-STING pathway, and this activation of this pathway in SMCs contributes to TAD progression^69,70^. Variants decreasing ENPP1 activity may therefore increase TAD risk by augmenting cGAS-STING signaling in SMCs.

***URM1*** functions in the post-translational urmylation of proteins together with its E1-activating enzyme UBA4 and also participates in the thiolation of wobble uridines in tRNA^71^. Thioredoxin peroxidase AHP1 detoxifies ROS to maintain redox homeostasis and is a target of URM1. Both AHP1 and tRNA thiolation contribute to URM1’s role in the oxidative stress response, and many TAD mouse models confirm that ROS levels increase in SMCs with TAD progression.

***CREM*** (cAMP Response Element Modulator) is structurally similar to the transcription factor CREB and can switch between activator and repressor states, thereby blocking CREB binding and fine-tuning cAMP signaling^72^. In the aorta, *CREM* expression is restricted to immune cells (Figure 4). However, a role for CREM in SMC phenotypic modulation is supported by the observation that *Crem*^-/-^ SMCs exhibit increased proliferation and *Crem*^-/-^ mice show increased neointimal formation following carotid artery injury^73^.

***SLFN11*** encodes a multifunctional protein that irreversibly blocks DNA replication under stress and inhibits viral replication^74^. SLFN11 has been extensively studied in cancer cells since its inactivation in 50% of cancer cell lines leads to chemoresistance. SLFN11-deficient cells have highlighted the protein’s diverse cellular functions, including roles in global ubiquitylation, endoplasmic reticulum stress, and protein aggregation^75^. SLFN11 also blocks mTOR signaling in hepatocellular carcinoma^76^, a signaling pathway shown in multiple mouse models to contribute to TAD pathogenesis^58^.

Unlike GWAS, burden analyses of rare variants in genes assess variants that disrupt the corresponding gene function, thereby providing a direct connection between the disrupted gene and TAD. Consequently, these analyses can be evaluated in terms of the possible mechanisms by which they cause the disease. HTAD genes harboring pathogenic variants that alter extracellular matrix proteins (*FBN1*, *LOX*) or SMC intracellular contractile proteins (*ACTA2*, *MYH11*, *MYLK*, *PRKG1*) disrupt the structure of the aortic medial layer termed the elastin-contractile unit. Disease-causing variants in these genes reduce force generation by aortic SMCs, thereby impairing mechanosensing. Two mechanosensing pathways implicated in TAD progression are YAP and focal adhesion kinase (FAK)^57,58^. YAP in SMCs protects the aorta from TAD progression, whereas FAK signaling drives both protective pathways, such as Rho/ROCK kinase signaling, and pathogenic pathways, including mTOR signaling. Thus, LOF and damaging missense rare variants in *ROCK1*, which is highly and selectively expressed in vascular SMCs, would be predicted to augment TAD . In contrast, mTOR inhibitors prevent TAD in multiple mouse models^58,77^, and since SLFN11 limits mTOR signaling, the loss of SLFN11 would potentially increase pathogenic mTOR signaling^76^. Downstream of these mechano-sensing pathways are stressed mitochondria, and CLEC16A regulates mitochondrial autophagy, which is critical for mitochondrial function and quality control. Stressed mitochondria increase ROS production and oxidative damage, and lowering ROS levels slows aneurysm growth in multiple TAD mouse models^78–80^. Thus, rare variants in *URM1* that prevent urmylation of AHP1 and thiolation of tRNA may increase ROS-driven cellular damage. Mitochondrial damage releases mitochondrial DNA into the cytosol, activating the cGAS-STING pathway, which has been shown to contribute to TAD progression^69,70^. ENPP1 hydrolyzes cGAMP, thus suppressing cGAS-STING activation, and loss of this suppression would promote TAD progression. Thus, the novel genes encode proteins involved in mechanosensing pathways, mitochondrial function, or signaling, and rare damaging missense variants in these genes are predicted to contribute to TAD by either disrupting protective pathways (e.g., *ROCK1*) or augmenting pathogenic pathways (e.g., *SLFN11*, *CLEC16A*, *URM1*, *ENPP1*).

Pathogenic variants in the genes for proteins involved in canonical TGFβ signaling also predispose to HTAD. Disease-causing variants lead to haploinsufficiency or missense variants that disrupt TGFβ signaling, implicating loss of canonical TGFβ signaling as a primary driver of TAD. *TGFBR1* and *TGFBR2* variants associated with early-onset and severe TAD also affect craniofacial development, leading to variable degrees of craniosynostosis, cleft palate, bifid uvula, and hypertelorism^29^. The function of *FNDC3B* is not clearly defined. However, loss of *FNDC3B* expression disrupts the TGFβ signaling network in multiple cell types and alters craniofacial development , and therefore may predispose to TAD in a manner similar to that of pathogenic variants in TGFBR1 and TGFBR2. While these craniofacial features were not observed in *FNDC3B* variant carriers in the biobanks, they have been described in individuals with microdeletions involving *FNDC3B*^53^.

Limitations of these results include that rare-variant burden testing is inherently constrained by the low frequency of ultra-rare alleles, which limits replication power and the precision of effect-size estimates. Consistent with this, some genes identified in the discovery analysis did not replicate in smaller biobank cohorts despite strong mechanistic plausibility, indicating that larger cohorts with genomic data will be required to resolve these signals. The independent replication cohorts were used to assess external support for the discovery-phase associations, while the subsequent five-biobank meta-analysis combined discovery and replication cohorts to maximize statistical power and estimate overall associations. Second, most of the participants analyzed were of European ancestry, limiting generalizability and underscoring the need for broader ancestral representation. We have also shown that a polygenic risk score for TAA modifies the penetrance of HTAD pathogenic variants in biobanks, and future work should integrate common and rare variants to refine risk prediction^81^. Larger sequencing cohorts will therefore be required to evaluate interactions between rare variants and potential modifiers of penetrance, including hypertension and polygenic risk.

If validated, these candidate genes could expand clinical genetic testing panels and improve the identification of individuals at increased risk for aortic dissection. As gene-specific risk increasingly informs surveillance and prophylactic surgery in HTAD^2^, defining the penetrance and natural history of these newly implicated genes may ultimately refine surgical thresholds and other individualized management strategies.

In summary, genome-wide aggregation of ultrarare, damaging missense and LOF variants across harmonized biobank datasets identifies established HTAD genes and novel candidate genes with large effect sizes. Because these associations involve protein-altering variants with predicted functional consequences, they offer more direct biological interpretability than common-variant GWAS findings and are broadly consistent with established TAD molecular pathways. Rare damaging variants in these novel candidate genes showed large-effect associations with TAD, with independent replication support and study-wide significance in the combined five-biobank meta-analysis, highlighting potential new contributors to clinically meaningful aortic disease risk. These findings refine the genetic architecture of TAD by identifying rare, high-risk variants in genes that may contribute to disease beyond familial HTAD. This study demonstrates a scalable framework for rare-variant burden testing in population-scale sequencing data and for uncovering high-impact genetic contributors to TAD and other genetically heterogeneous cardiovascular conditions.

## Data Availability

The UKB and AoU data are available to researchers upon approval by their respective access committees. The UTHealth datasets are available in dbGaP Study Accession: phs000693.v7.p3. The PMBB, MGBB, and MVP datasets are not publicly available due to IRB restrictions requiring a collaboration with an associated investigator.

## Acknowledgements

We gratefully acknowledge the participants of the UK Biobank, All of Us Research Program, Penn Medicine BioBank, Mass General Brigham Biobank, and VA Million Veteran Program, without whom this research would not have been possible. A large language model (ChatGPT, OpenAI) was used to assist with language editing and improving readability and with the preparation of the study design schematic shown in Figure 1. All scientific content and interpretations were provided and verified by the authors.

## Non-standard Abbreviations and Acronyms

AoU: All of Us Research Program
ESTAD: Early-Onset Sporadic Thoracic Aortic Dissection
GWAS: genome-wide association study
HTAD: heritable thoracic aortic disease
LOF: loss-of-function
MAF: minor allele frequency
MGBB: Mass General Brigham Biobank
MVP: Million Veteran Program
PMBB: Penn Medicine Biobank
PV: pathogenic variant
scRNA-seq: single-cell RNA sequencing
SMC: smooth muscle cell
TAA: thoracic aortic aneurysm
TAD: thoracic aortic disease
UKB: UK Biobank.

