## Supplemental Material for "Gene-Based Rare Variant Burden Analyses Across Biobanks Identify Novel High-Risk Genes for Thoracic Aortic Disease"

**Supplemental Methods**

**Biobank Cohorts**

The UK Biobank (UKB) is a population-based study of approximately 500,000 individuals recruited across the United Kingdom^17^. The research presented here was conducted under UKB Application Number 75470 using data from release v18.1 (November 2023). We included genome sequencing data from participants of European ancestry, defined by self-report as “British,” “Irish,” “White,” or “Any other White background,” comprising approximately 95% of the UKB cohort.

The All of Us (AoU) Research Program is a large-scale initiative that collects health and genomic data from individuals across the United States^18^. For this study, we used the AoU v8 dataset, which includes genome sequencing data from >414,000 participants of diverse ancestral backgrounds.

To replicate our primary meta-analysis findings, we analyzed exome sequencing data from 41,888 participants in the Penn Medicine Biobank (PMBB) and 9,406 participants in the Mass General Brigham Biobank (MGBB), along with genome sequencing data from 77,122 participants in the VA Million Veteran Program (MVP)^4,19,20^. Complete biobank demographic data are provided in Table S1.

The PMBB is a genomic and precision medicine cohort comprising Penn Medicine health system patients who consent to linkage of electronic health records with biospecimens, including 43,731 participants who have undergone ES^19^. The PMBB is approved under University of Pennsylvania IRB protocol #813913. The MGBB is a hospital-based biobank comprising Mass General Brigham patients who consent to linkage of biospecimens and genomic data with electronic health record data^20^. The MVP is a large genomic and precision medicine cohort of U.S. Veterans recruited from participating Department of Veterans Affairs (VA) medical facilities^4^. Participants provide written informed consent for research participation and linkage of genomic data with longitudinal electronic health record data. The MVP received ethical and study protocol approval from the VA Central Institutional Review Board.

**Disclosure Control and Reporting of Participant Counts**

Reporting of participant counts was subject to each biobank’s disclosure-control policies. UKB requires minimum aggregate counts for public reporting, while AoU prohibits publication of counts from 1–20 or statistics from which such counts could be derived. Other contributing biobanks impose similar restrictions on reporting small participant counts. Accordingly, cohort-specific carrier counts, case-control carrier breakdowns, and allele-level summaries were reported only where permitted.

**Additional Clinically Ascertained Cohorts**

***UTHealth Heritable Thoracic Aortic Disease (HTAD) Cohort***
 The UTHealth HTAD cohort included 437 probands from unrelated families with heritable thoracic aortic disease, defined as families with ≥2 affected members with thoracic aortic aneurysm or dissection. Exome or genome sequencing was performed on blood-derived DNA.

***Early-Onset Sporadic Thoracic Aortic Dissection (ESTAD) Cohort***
 The ESTAD cohort comprised 551 individuals aged ≤60 years presenting with thoracic aortic dissection without syndromic features or a known family history of aortic disease. Clinical data were reviewed to exclude individuals meeting criteria for recognized syndromic aortopathies. Exome or genome sequencing was performed on blood-derived DNA.

***Yale Aortic Surgery Cohort***
 The Yale cohort included 1,750 adults undergoing emergent surgery for acute thoracic aortic dissection or elective repair of thoracic aortic aneurysm. Exome sequencing was performed on DNA extracted from excised aortic tissue. Clinical phenotypes were abstracted from operative and medical records.

***GeneDx Trio***
 A clinically ascertained family trio was identified through physician-ordered exome sequencing and contributed by GeneDx under a research protocol approved by the Western Institutional Review Board (Study Number 1169768), which met criteria for a waiver of informed consent.

**Sequencing Data Processing, Quality Control, and Variant Annotation**

Sequencing data from the UKB, consisting of multi-sample project variant call format (pVCF) files generated with the Illumina DRAGEN pipeline (v3.7.8, aligned to GRCh38), were accessed via the UKB Research Analysis Platform (RAP) and processed using custom Python scripts within that environment. Sequencing data from the AoU cohort, also processed with DRAGEN v3.7.8, were accessed via Hail within the AoU Researcher Workbench. Variant-level filters for both UKB and AoU included DRAGEN PASS status, QUAL ≥ 30, and <10% missingness. Genotype-level filters required read depth (DP) ≥ 10, genotype quality (GQ) ≥ 20, allele balance ≥ 0.2 for heterozygous genotypes, and exclusion of variants with excess heterozygosity. Sequencing and quality control procedures for PMBB, MGBB, and MVP have been described previously^4,19,20^**.**

Variant annotation and functional effect prediction were performed using Ensembl comprehensive protein-coding transcripts, and results were summarized based on MANE Select transcripts^82,83^. For the UKB cohort, we used bcftools, SnpEff, and SnpSift to annotate and extract qualifying variants from project VCF (pVCF) files^84–86^. Annotations included minor allele frequencies (MAF) from gnomAD v4 and pathogenicity predictions from dbNSFP v4.8^14,87^. Identical filtering criteria were applied to the AoU cohort using pre-annotated Hail tables generated by Nirvana v3.18, with gnomAD MAF annotations provided through Nirvana and missense pathogenicity predictions from dbNSFP^88^. Variants in the three replication cohorts (PMBB, MGBB, and MVP) were identified using analogous pipelines and identical filtering thresholds.

**Derivation of Rare Missense Variant Filtering Thresholds**

The gnomAD MAF, REVEL, and AlphaMissense thresholds used for missense variant burden testing were derived previously using a data-driven optimization procedure in the PMBB^13–16^ Combinations of allele-frequency and missense pathogenicity thresholds were evaluated for their ability to distinguish variants associated with aortic dissection, with the optimal cutpoints selected by maximizing the Youden index. Stability of the selected thresholds was assessed by bootstrap validation, and the resulting parameters were subsequently evaluated in independent cohorts. This procedure yielded thresholds of gnomAD MAF <8.16 × 10⁻⁶, REVEL >0.649, and AlphaMissense >0.2543, which were prespecified and applied unchanged in the current genome-wide burden analyses.

**Sex-Stratified and Gene-by-Sex Interaction Analyses**

For the 14 genes meeting the prespecified cross-cohort validation criteria, sex-stratified burden analyses were performed separately in females and males in UKB and AoU using the same qualifying variant definitions and Firth penalized logistic regression framework as the primary analyses. Models included age and the first four genetic principal components in UKB or the first five in AoU. Sex was omitted as a covariate in the sex-stratified analyses. Analyses were limited to UKB and AoU because sparse qualifying variant carrier counts in the replication cohorts limited reliable sex-specific effect estimation.

Gene-by-sex interaction was evaluated in each cohort by including rare variant carrier status, sex, and a carrier status-by-sex interaction term in the regression model, along with age and genetic principal components. Sex was coded as female = 0 and male = 1; therefore, the interaction OR represents the ratio of the variant-associated OR in males relative to females, with values >1 indicating a stronger association in males and values <1 indicating a stronger association in females. Female-specific, male-specific, and gene-by-sex interaction effect estimates were meta-analyzed separately across UKB and AoU using fixed-effect inverse-variance weighting. Meta-analysis was limited to effects available in both cohorts. If a sex had no case carriers in a given cohort, the corresponding sex-specific estimate was reported as unavailable; if either sex had no case carriers, the gene-by-sex interaction estimate for that cohort was also reported as unavailable. Interaction P values were considered exploratory and were not adjusted for multiple testing.

**Cell type prioritization in scRNA-seq data**

Previously published thoracic aorta scRNA-seq datasets from control (n=5 donors),

aneurysm (n=8), and dissection (n=6) samples were reanalyzed^36,37^. Raw FASTQ files were processed with Cell Ranger (v9.0.1) using the GRCh38 reference (GENCODE v44/Ensembl 110 annotation). Spliced and unspliced reads were quantified with velocyto (v0.17.17)^89^. Ambient RNA was removed using CellBender (v0.3.2)^90^. Only droplets detected in the Cell Ranger filtered matrix and classified as non-empty with probability >0.99 by CellBender were retained to minimize false-positive droplets. Low-quality cells were removed based on the following thresholds: background fraction ≥15%; mitochondrial RNA fraction ≥30%; extreme unspliced-to-spliced ratios (<0.1% or >10%); ≤250 genes; ≤650 UMIs, or Gini coefficient ≥0.8. Thresholds were selected based on empirical QC distributions and prior single-cell studies.

Doublets were removed using a consensus approach. We applied DoubletFinder (v2.0.6)^91^ with (and without) reference labels, scds (v1.22)^92^ hybrid cxds-bcds scoring, and scDblFinder (v1.20.2)^93^ with (and without) labels. The expected doublet rate in DoubletFinder was set to follow 10x Genomics guidelines. Cells flagged as doublets by at least three of the five configurations were excluded. Additional doublets identified during clustering and annotation stage were also removed.

Raw counts were log-normalized in Seurat (v5.3.1)^94^ with a scale factor of 10,000. Variable genes were selected using VST (2,000 genes). Each dataset underwent independent PCA, and reciprocal anchors were identified using the top 30 PCs. RPCA integration in Seurat was chosen to reduce over-correction and preserve donor-level variation. Clustering was performed using the Leiden algorithm on a nearest-neighbor graph; resolution was tuned to stabilize rare populations and to help identify potential low QC cells. UMAP embeddings were computed using 30 neighbors, with default settings. Clusters were annotated using canonical markers, differential markers, and QC features. Clusters with aberrant QC metrics unexplained by biological identity or enriched for doublets were removed. Meta-analysis-validated genes were queried in the integrated dataset.

**Supplemental Figures**

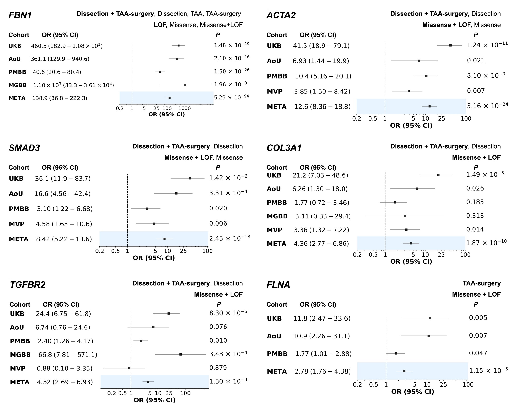

**Figure S1. Association of established and putative HTAD genes across discovery and replication cohorts.**

Cohort-specific and fixed-effect inverse-variance weighted meta-analytic odds ratios (ORs) with 95% confidence intervals and *P* values are shown for the **five established and one putative HTAD genes** that met the cross-cohort validation criteria. Replication cohorts without any case carriers for a given gene were excluded. For each gene, the TAD phenotypes and variant classes (missense, LOF, or missense+LOF) that reached study-wide significance in the UKB + AoU discovery analysis are indicated. The most significant phenotype–variant class combination carried forward into replication is indicated in **bold**. UKB, UK Biobank; AoU, All of Us; PMBB, Penn Medicine Biobank; MGBB, Mass General Brigham Biobank; MVP, Million Veteran Program.

**VA Million Veteran Program**

**MVP Program Office**

- Sumitra Muralidhar, Ph.D., Program Director

US Department of Veterans Affairs, 810 Vermont Avenue NW, Washington, DC 20420

- Jennifer Moser, Ph.D., Associate Director, Scientific Programs

US Department of Veterans Affairs, 810 Vermont Avenue NW, Washington, DC 20420

- Jennifer E. Deen, B.S., Associate Director, Cohort & Public Relations

US Department of Veterans Affairs, 810 Vermont Avenue NW, Washington, DC 20420

**MVP Steering Committee**

- Co-Chair: J. Michael Gaziano, M.D., M.P.H.

VA Boston Healthcare System, 150 S. Huntington Avenue, Boston, MA 02130

- Co-Chair: Dave Oslin, M.D.

Philadelphia VA Medical Center, 3900 Woodland Avenue, Philadelphia, PA 19104

- Sumitra Muralidhar, Ph.D., Ex-Officio

US Department of Veterans Affairs, 810 Vermont Avenue NW, Washington, DC 20420

- Drew Helmer, M.D., M.S.

Michael E. DeBakey VA Medical Center, 2002 Holcombe Boulevard, Houston, TX 77030

- Adriana Hung, M.D., M.P.H.

VA Tennessee Valley Healthcare System, 1310 24th Avenue, South Nashville, TN 37212

- Philip S. Tsao, Ph.D.

VA Palo Alto Health Care System, 3801 Miranda Avenue, Palo Alto, CA 94304

- Deepak Voora, M.D.

Durham VA Medical Center, 508 Fulton Street, Durham, NC 27705

**MVP Co-Principal Investigators**

- J. Michael Gaziano, M.D., M.P.H.

VA Boston Healthcare System, 150 S. Huntington Avenue, Boston, MA 02130

- Philip S. Tsao, Ph.D.

VA Palo Alto Health Care System, 3801 Miranda Avenue, Palo Alto, CA 94304

**MVP Core Operations**

- Jessica V. Brewer, M.P.H., Director, MVP Cohort Operations

VA Boston Healthcare System, 150 S. Huntington Avenue, Boston, MA 02130

- Kelly Cho, M.P.H., Ph.D., Director, MVP Phenomics

VA Boston Healthcare System, 150 S. Huntington Avenue, Boston, MA 02130

- Lori Churby, B.S., Director, MVP Regulatory Affairs

VA Palo Alto Health Care System, 3801 Miranda Avenue, Palo Alto, CA 94304

- Yonghui Jia, Ph.D., Director, VA Central Biorepository

VA Boston Healthcare System, 150 S. Huntington Avenue, Boston, MA 02130

- Jacob T. Kean, Ph.D., Acting Director, VA Informatics and Computing Infrastructure (VINCI)

VA Salt Lake City Health Care System, 500 Foothill Drive, Salt Lake City, UT 84148

- Saiju Pyarajan, Ph.D., Director, Data and Computational Sciences

VA Boston Healthcare System, 150 S. Huntington Avenue, Boston, MA 02130

- Robert Ringer, Pharm.D., Director, VA Albuquerque Central Biorepository

New Mexico VA Health Care System, 1501 San Pedro Drive SE, Albuquerque, NM 87108

- Luis E. Selva, Ph.D., Director, MVP Biorepository Coordination

VA Boston Healthcare System, 150 S. Huntington Avenue, Boston, MA 02130

- Shahpoor (Alex) Shayan, M.S., Director, MVP PRE Informatics

VA Boston Healthcare System, 150 S. Huntington Avenue, Boston, MA 02130

- Brady Stephens, M.S., Principal Investigator, MVP Information Center

Canandaigua VA Medical Center, 400 Fort Hill Avenue, Canandaigua, NY 14424

- Stacey B. Whitbourne, Ph.D., Director, MVP Cohort Development and Management

VA Boston Healthcare System, 150 S. Huntington Avenue, Boston, MA 02130

### **Penn Medicine BioBank**

#### **PMBB Leadership Team**

Daniel J. Rader, M.D., Marylyn D. Ritchie, Ph.D.

**Contribution**: All authors contributed to securing funding, study design and oversight. All authors reviewed the final version of the manuscript.

#### **Patient Recruitment and Regulatory Oversight**

JoEllen Weaver, Nawar Naseer, Ph.D., M.P.H., Giorgio Sirugo, M.D., Ph.D., Afiya Poindexter, Jenna Dever, Aidan Harvey, Sydney Linn, Naman Srivastava

**Contributions**: JW manages patient recruitment and regulatory oversight of study. NN manages participant engagement, assists with regulatory oversight, and researcher access. GS assists with researcher access. AP, JD, AH, SL, and NS perform recruitment and enrollment of study participants.

#### **Lab Operations**

JoEllen Weaver, Meghan Livingstone, Fred Vadivieso, Stephanie DerOhannessian, Teo Tran, Julia Stephanowski, Salma Santos, Ned Haubein, Ph.D., Joseph Dunn

**Contribution**: JW, ML, FV, SD conduct oversight of lab operations. ML, FV, SD, TT, JS, SS perform sample processing. NH, JD are responsible for sample tracking and the laboratory information management system.

#### **Clinical Informatics**

Anurag Verma, Ph.D., Colleen Morse Kripke, M.S. DPT, MSA, Marjorie Risman, M.S., Renae Judy, B.S., Colin Wollack, M.S.

**Contribution**: All authors contributed to the development and validation of clinical phenotypes used to identify study subjects and (when applicable) controls.

#### **Genome Informatics**

Anurag Verma Ph.D., Shefali S. Verma, Ph.D., Scott Damrauer, M.D., Yuki Bradford, M.S., Scott Dudek, M.S., Theodore Drivas, M.D., Ph.D., Zachary Rodriguez, Ph.D,

**Contribution**: AV, SSV, and SD are responsible for the analysis, design, and infrastructure needed to quality control genotype and exome data. YB performs the analysis. TD and AV provide variant and gene annotations and their functional interpretation of variants.
